# The cross-sectional area of the vagus nerve is not reduced in Parkinson’s Disease patients

**DOI:** 10.1101/2020.10.19.20214973

**Authors:** Laura C.J. Sijben, Werner H. Mess, Uwe Walter, A. Miranda L. Janssen, Mark Kuijf, Mayke Oosterloo, Wim M. Weber, Marcus L.F. Janssen

**Affiliations:** Department of Clinical Neurophysiology, Maastricht University Medical Center, Maastricht, The Netherlands; School for Mental Health and Neuroscience, Faculty of Health, Medicine and Life Sciences, Maastricht University, Maastricht, The Netherlands; Department of Neurology, University of Rostock, Rostock, Germany; German Centre for Neurodegenerative Diseases (DZNE), Rostock, Germany; Department of Methodology and Statistics, School for Public Health and Primary Care, Maastricht University; Department of Neurology, Maastricht University Medical Center, Maastricht, The Netherlands

**Keywords:** Parkinson’s Disease, vagus nerve, ultrasonography, cross-sectional area, autonomic symptoms

## Abstract

**Objective:** Recent studies have revealed the importance of the gut brain axis in the development of Parkinson’s disease (PD). It has also been suggested that the cross-sectional area (CSA) of the vagus nerve can be used in the diagnosis of PD. Here, we hypothesize that the CSA of the vagus nerve is decreased in PD patients compared to control subjects.

**Methods:** In this study we measured the CSA of the vagus nerve on both sides in 31 patients with PD and 51 healthy controls at the level of the common carotid artery using high-resolution ultrasound.

**Results:** The CSA of the vagus nerve was not reduced in PD patients compared to controls (p = 0.391. The mean CSA of the left vagus nerve was significantly smaller than the right (p < 0.001). There was no significant correlation between age, gender and autonomic symptoms with the CSA of the left (p = 0.128) and right vagus nerve (p = 0.166).

**Conclusion:** These findings show that the CSA of the vagus nerve using ultrasonography is not a reliable diagnostic tool in the diagnosis of PD.

**Highlights:**

- The cross-sectional area of the vagus nerve is not decreased in Parkinson disease patients.
- The cross-sectional area of the vagus nerve does not correlate with autonomic symptoms.
- Measurement of the vagus nerve cross-sectional area has a high inter-observer correlation.

## 1. Introduction

Parkinson’s disease (PD) is a progressive neurodegenerative disorder resulting in a clinical hypokinetic rigid syndrome. Besides the classical motor symptoms, PD is associated with non-motor symptoms. Among these are autonomic symptoms, such as orthostatic hypotension, constipation and urinary incontinence. Autonomic symptoms are frequently reported by PD patients and have been suggested as preclinical symptoms [1].

The main neuropathologic hallmark is the formation of cytoplasmic inclusions called α-synuclein-enriched Lewy bodies and Lewy neurites. These α-synuclein aggregations are found throughout the brain, most pronounced in the substantia nigra [2, 3]. Neural cell loss is most abundant in the substantia nigra compacta. Interestingly, the pathological α-synuclein is also present in the peripheral nervous system, more specifically in the vagus nerve, which plays an important role in autonomic control [4, 5]. Braak and colleagues hypothesized that a neurotropic pathogen entering the brain may be involved in the pathology of PD [5, 6]. This may occur via nasal or gastric route, causing transsynaptic cell-to-cell, prion like, transmission of α-synuclein [5, 6]. It is suggested that the pathogen invades the brain by entering the nasal mucosa, followed by the olfactory bulb and the anterior olfactory nucleus into the olfactory structures of the temporal lobe. The gastric route starts in the enteric nervous system, where α-synuclein enters the Meissner’s and Auerbach’s plexus. From that point α-synuclein is transported to the lower part of the brainstem through the vagus nerve. α-synuclein spreads from the vagus dorsal motor nucleus into the medulla and the rest of the brain [6]. A recent study in an animal model supports the hypothesis that the propagation of pathologic α-synuclein via vagus nerve causes PD [7].

In line with these findings it has been shown that a vagotomy or appendectomy early in life are associated with a lower risk to develop PD later in life [8-10] [11-14]. The appendix appears to be the largest source of α-synuclein, whereas the vagus nerve only seems to be the transporter [14, 15]. Based on these findings it has been postulated that the cross-sectional area (CSA) of the vagus nerve decreases in PD. In fact, several studies have been published showing a significant decrease in the CSA of the vagus nerve in PD [16-18]. It has even been hypothesized that atrophy of the vagus nerve already occurs at a pre-clinical stage [16]. These findings highlight the potential to use the assessment of the CSA of the vagus nerve rostral to the carotid bifurcation by using B-mode ultrasound as a fast and low-cost diagnostic test. Moreover, this method potentially identifies persons at risk even before a clinical diagnosis of PD can be confirmed. It is however important to note that two studies using B-mode ultrasound could not confirm that the CSA of the vagus nerve was decreased in PD [19, 20].

The primary aim of this study is to assess if the CSA is decreased in PD. We hypothesize that the CSA of the vagus nerve is reduced in PD patients compared to controls using ultrasonography. Our secondary aim is to assess if the CSA correlates with autonomic symptoms in PD. We will also assess the interrater reliability and compare two different methods to determine the CSA. To these aims, we conducted a cross-sectional study in which we acquired the CSA of the left and right vagus nerve using ultrasonography. Autonomic symptoms were assessed by a standardized questionnaire.

## 2. Material and methods

The study was given a positive advice by the local ethics committee and all participants gave written informed consent before participation (*METC Maastricht University Medical Center, the Netherlands, protocol number 2019-1223*). All participants underwent a B-mode ultrasound examination of the vagus nerve and filled in a short questionnaire about autonomic complaints (Scales for Outcomes in Parkinson’s Disease - Autonomic Dysfunction; SCOPA-AUT).

### 2.1 Subjects

The inclusion criterion was a clinical diagnosis of PD made by the treating neurologist in accordance with the UK Brain Bank criteria for the PD group [21]. The control group consisted of persons without PD. Exclusion criteria for both groups were other neurodegenerative disorder(s) and previous surgery in proximity of the vagus nerve (for example carotid endarterectomy or implantation of vagus nerve stimulator). Participants were recruited by their neurologist. Recruitment of controls was performed at the clinic in patients whose carotid arteries were examined since a transient ischemic attack was suspected.

### 2.2 Ultrasound

Ultrasound of the vagus nerve was performed by experienced nerve sonographers using the Philips IU22 with a linear 17.5 MHz transducer. The sonographer was not blinded for the subject’s identity. The vagus nerve was visualized in a coronal plane anterolateral to the common carotid artery (Figure 1). To calculate the CSA of the vagus nerve, the contour of the vagus nerve within the hyperechoic epineural rim was outlined. The Philips IU22 then provides the CSA automatically. At each side the CSA of the vagus nerve was measured twice. The mean of the two measurements was used for statistical analyses.

**Figure 1.**
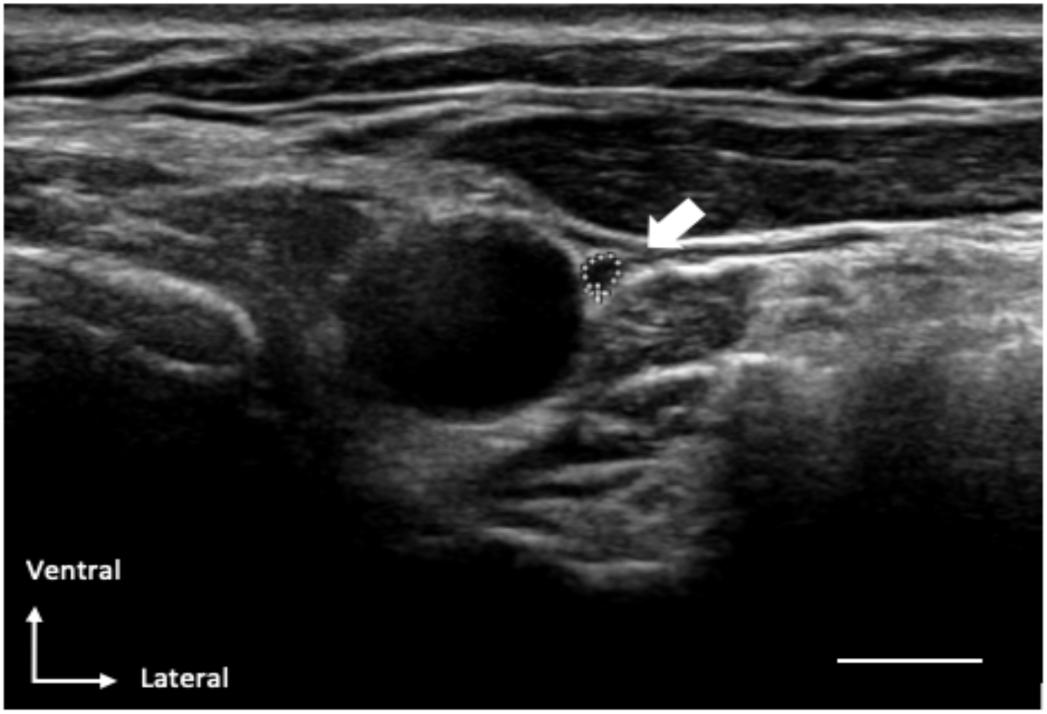
Ultrasound image. Representative example of an ultrasound image in which the vagus nerve (dotted circle) is positioned ventrolateral to the common carotid artery. The scale bar equals 0.5 cm.

To ensure a good interrater reliability, the contour of the vagus nerve within the hyperechoic epineural rim was outlined by two independent sonographers on the acquired ultrasound images in ten participants.

To asses possible differences between our method applied (as described above) and one of the earlier studies [18], we measured the longest cross-sectional diameter *a* and the diameter *b* perpendicular to *a* (as described by Walter et al.) in 34 participants. The CSA was calculated using the following formula: *(a*b*π)/4*. The diameters were measured at the same place as the contour of the vagus nerve was measured.

### 2.3 Questionnaire

On the same day as the ultrasound examination took place all participants filled in the SCOPA-AUT, a validated questionnaire about autonomic complaints [22, 23]. The SCOPA-AUT consists of six domains: gastrointestinal, urinary, cardiovascular, thermoregulatory, pupillomotor and sexual functioning. There are different numbers of questions for each domain, with a total of 23 questions. For each question a score from 0-3 points can be obtained, the maximum score that can be obtained is 69 points. The total sum score was used for further analyses.

### 2.4 Statistics analysis

Baseline characteristics from the PD and control group were compared with an independent-samples t-test for age and the result on the SCOPA-AUT and with a Chi-square test for gender. As a first step of the statistical analysis, a repeated measures ANOVA (with side as within-subject factor and group as between subject factor) to test for possible differences between the left and right CSA, and differences between groups was performed. Secondly, a linear regression analysis was conducted using clinical and demographic variables (gender, age, group and result on the SCOPA-AUT) as predictors to test their effect on the CSA of the vagus nerve. The linear regression was conducted for the CSA of the vagus nerve of the left and right side.

The Intraclass Correlation Coefficient (ICC(2,1) classification according to Shrout and Fleiss) was calculated to compare the two different types of measurements for the CSA and to compare the observations by the two independent sonographers [24]. The SPSS statistical computer package (version IBM SPSS Statistics for Macintosh, version 25 (IBM Corp., Armonk, N.Y., USA)) was used for all statistical analyses.

## 3. Results

### 3.1 Demographics

We recruited 82 participants (31 PD and 51 controls) between December 2019 and April 2020 at the Departments of Neurology and Clinical Neurophysiology of Maastricht University Medical Center (MUMC+). The clinical and ultrasonographic findings are summarized in table 1.

**Table 1.** Demographics, clinical scores and ultrasonography of the Parkinson and control group. Values except gender are presented as means + SD. For gender number and percentage of females are given. ^a^P-values are from comparisons between PD patients and the control group using an independent-samples t-tests for age and SCOPA-AUT and a repeated measurement ANOVA for the CSA of the left and right vagus nerve. CSA, cross-sectional area in mm^2^. SCOPA-AUT, Scales for Outcomes in Parkinson’s Disease - Autonomic Dysfunction.

|  | *Parkinson (n=31)* | *Control (n=51)* | *P<sup>a</sup>* |
| --- | --- | --- | --- |
| **Demographics** |  |  |  |
| *Mean age (years)* | 69 (8) | 72 (8) | 0.262 |
| *Gender (female)* | 15 (48%) | 22 (43%) | 0.643 |
| *Disease duration (months)* | 95 (66) |  |  |
| **Clinical scores** |  |  |  |
| *SCOPA-AUT* | 16 (7) | 13 (8) | 0.078 |
| **Ultrasonography (CSA, mm<sup>2</sup>)** |  |  |  |
| *Left vagus nerve* | 2.10 (0.57) | 1.90 (0.56) | 0.391 |
| *Right vagus nerve* | 2.54 (0.70) | 2.24 (0.68) | 0.391 |

In the control group, nine (18%) participants had diabetes mellitus without complications, 31 (62%) participants had cardiovascular disease, and one (2%) had a polyneuropathy. The average number of medications the participants used at the time of the ultrasound was four (range: 0-11). In one participant the medical history was unavailable. In the Parkinson group, two (7%) patients had diabetes mellitus without complications, five (16%) patients had cardiovascular disease and none of the participants had a history of polyneuropathy. The average number of medications the participants used apart from their Parkinson medication at the time of the ultrasound was two (range: 0-14).

### 3.2 CSA of the vagus nerve

First, we assessed if a good interrater reliability was achieved (figure 2). The ICC showed a strong agreement between the independent sonographers (ICC = 0.969, p < 0.001.). Secondly, we assessed if our investigation method was compatible with an earlier described method to estimate the CSA using the diameter of the vagal nerve in two directions [18]. The ICC showed a strong agreement between the two methods to estimate the CSA (ICC = 0.961, p < 0.001).

**Figure 2.**
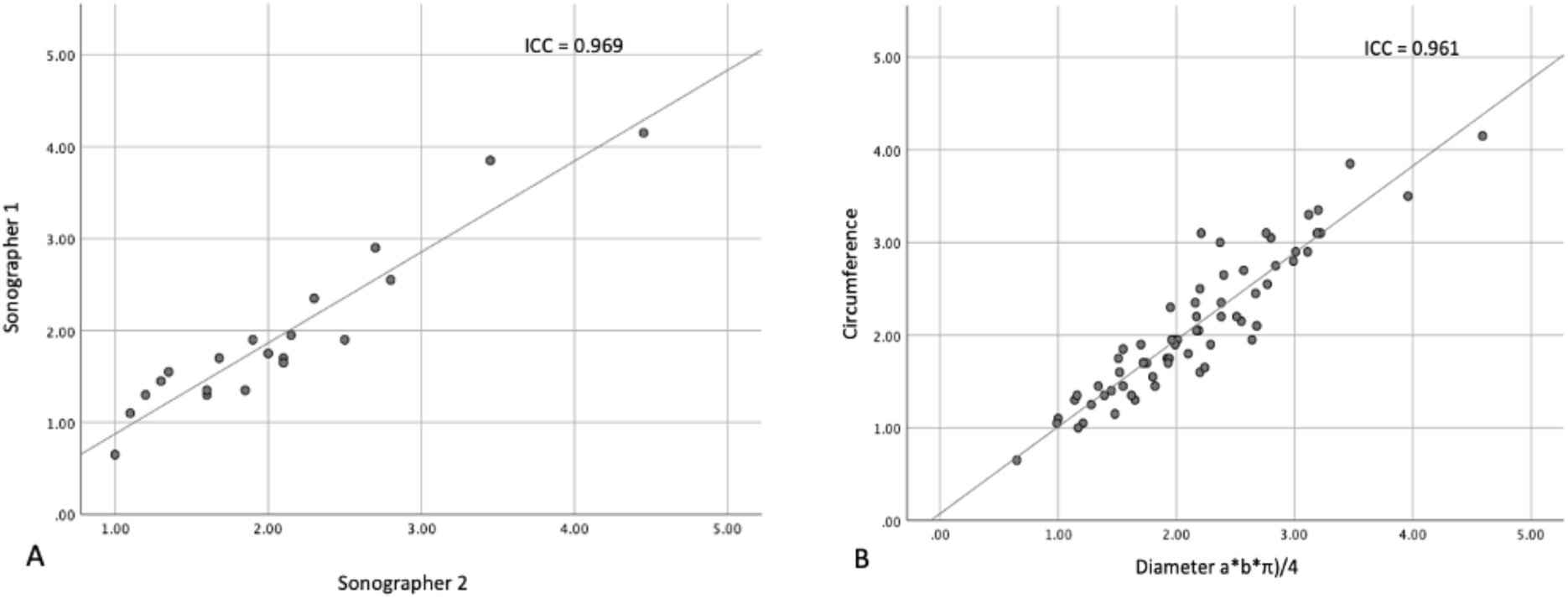
Scatterplots showing the linear correlation between A) the CSA determent by two independent sonographers, B) the different methods to obtain the CSA (respectively circumference on the y- and diameter a*b*π)/4 on the x-axis).

The mean CSA of the left vagus nerve in the PD and the control group was respectively 2.10 ± 0.57 mm^2^ and 1.90 ± 0.56 mm^2^ and of the right respectively 2.54 ± 0.70 mm^2^ and 2.24 ± 0.68 mm^2^ (figure 3). The repeated measures ANOVA determined that the mean CSA of both the left and right vagus nerve did not differ statistically between the PD and control group (F(1,80) = 0.745, p = 0.391). Furthermore, the repeated measures ANOVA determined that the mean CSA of the left vagus nerve was significantly smaller than the right in both PD and controls (F(1,80) = 42.864, p < 0.001). The difference between left and right vagus nerve CSA was 0.391 (95%-CI [0.273, 0.511]) mm^2^.

**Figure 3.**
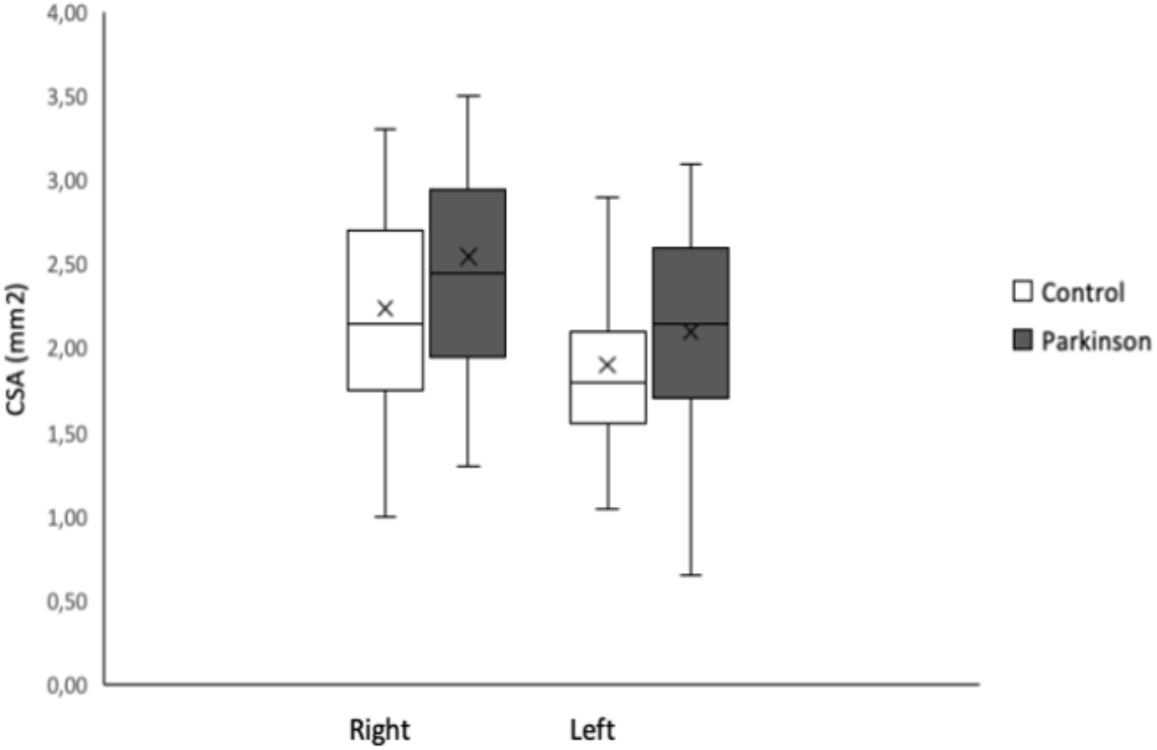
Barplots representing the mean of the CSA of the left and right vagus nerve of the Parkinson Disease and control group. The mean is shown with an “X” and the median with a horizontal line. Error bars represent standard deviation of the mean. CSA, cross-sectional area in mm^2^.

### 3.3 Correlation between autonomic symptoms and CSA of the vagus nerve

The mean total score on the SCOPA-AUT was 16 ± 7 in the PD and 13 ± 8 points in the control group. There was no significant difference between the total SCOPA-AUT score of the two groups (p = 0.078). Group, age and gender did not show a significant correlation with the CSA of the left vagus nerve (F(4, 77) = 1.851, p = 0.128 with an R^2^ of 0.088) or the right vagus nerve (F(4, 77) = 1.666, p = 0.166 with an R^2^ of 0.080).

## 4. Discussion

In this cross-sectional study, we show that the CSA of the vagus nerve is not reduced in PD patients compared to controls. In addition, no relation between the CSA of the vagus nerve and autonomic symptoms as assessed by the SCOPA-AUT questionnaire was found. The CSA was neither influenced by gender or age. Consistent with previous studies, the CSA of the right vagus nerve was significantly larger in both the PD and control group [16-19, 25, 26].

Our findings confirm the findings of two studies which also found no reduction in CSA of the vagus nerve in PD patients [19, 20]. We were unable to replicate the reduction in CSA as reported by earlier studies [16-18]. Important to note is the high variation in CSA measured between studies. The mean CSA in control subjects varies from 1.3 to 2.7 mm^2^ for the right and 1.1 to 2.6 for the left vagus nerve. In the PD groups the variation is even larger between studies. In our cohort we also found a high variance in CSA between individuals.

The validity of our findings is supported by several factors. First, we have shown a high inter-rater reliability. Second, our method (drawing a circle within the epineural rim to determine the CSA) has a high correlation with previously used method, in which the longest cross-sectional diameter *a* and the diameter *b* perpendicular to *a* were used to calculate the CSA [18]. Third, our findings replicate the well-known left – right difference of the CSA of the vagus nerve [16-19, 25, 26]. This left-right difference in CSA is explained by the different functional anatomy of the vagus nerve. The right vagus nerve innervates parts of the small intestine, the colon and also a part of the gastric plexus. The left vagus nuclei terminate in the anterior plexus and from there on new branches go to the stomach, liver and the superior part of the abdomen [16, 27]. Fourth, the range of the CSA of the vagus nerve we found is similar to previous reports [16, 19, 25].

A possible reason why in our study no difference was found between PD and controls is the low prevalence of autonomic symptoms in our PD group. A typical mean score at the SCOPA-AUT for Parkinson patients is normally around 25 points [22]. It could be postulated that patients with more autonomic symptoms might show a reduced CSA of the vagus nerve. On the other hand, the linear regression analysis did not show any relation between the SCOPA-AUT scores and the CSA of the vagus nerve. One could argue that our control group was not a completely healthy population. The control group was recruited from a group with a higher risk on cardiovascular comorbidities. A reliable diagnostic tool to diagnose PD should however also be able to differentiate between patients suffering from PD and person with a higher risk on cardiovascular complications. A limitation of our study is that the sonographer was not blinded for the diagnosis. We argue that this has not limited the objective assessment of the epineural rim and defining the CSA of the vagus nerve in our cohort. Especially, since we used state of the art high-resolution ultrasonography, with a 17 MHz transducer which is higher than the probes used in earlier studies. In thus remains a challenging search to find a reliable and cheap biomarker in the early or preclinical diagnosis of PD. Considering the high interindividual variance the CSA and the low interrater reliability. A reduction over time per individual might be able to provide a better prediction on the risk of developing PD.

## 5. Conclusion

The CSA of the vagus nerve can be measured with high accuracy. It is not reduced in PD patients and is neither correlated with autonomic symptoms, gender nor age. Therefore, we conclude that a single ultrasound examination of the vagus nerve is at this moment not a suitable biomarker in the diagnosis of PD.

## Data Availability

All data can be requested by email to the corresponding author.

## 6. Acknowledgements

We would like to thank Nathal van Golde and Chantal Ruppe for their technical assistance.

